# Quantifying the Regional Disproportionality of COVID-19 Spread

**DOI:** 10.1101/2024.04.05.24305413

**Authors:** Kenji Sasaki, Yoichi Ikeda, Takashi Nakano

**Affiliations:** Center for Infectious Disease Education and Research, Osaka University, Osaka 565-0871, Japan; Research Center for Nuclear Physics, Osaka University, Osaka 567-0047, Japan

**Keywords:** Infectious disease, COVID-19, Epidemiology, Pandemic, Inequality measure, Information theory, Kullback-Leibler divergence

## Abstract

**Background:** The COVID-19 pandemic has caused serious health problems and has had major economic and social consequences worldwide. Understanding how infectious diseases spread can help mitigating the social and economic impact.

**Objective:** The study focuses to capture the degrees of disproportionality in prevalence rates of infectious disease across different regions over time.

**Methods:** We analyze the numbers of daily COVID-19 confirmed cases in the United States collected by Johns Hopkins University over 1100 days since the first reported case in January 2020 in order to assess quantitatively the disproportionality of the confirmed cases using the Theil index, a measure of imbalance used in economics. Results:

Our results reveal a dynamic pattern of interregional disproportionality in the confirmed cases by monitoring variations in regional contributions to the Theil index as the pandemic progresses.

**Conclusions:** The combined monitoring of this indicator and the confirmed cases is crucial for understanding regional differences in infectious diseases and for effective planning of response and resource allocation.

## Introduction

The COVID-19 pandemic has caused serious health problems and has had major economic and social consequences worldwide. A number of indicators and models have been proposed to address the problem, and mechanisms for the spread of the infection and intervention measures to control the pandemic have been studied [1– 7].

Several studies have investigated regional differences in COVID-19 prevalence [8– 11]. Differences in prevalence rates between regions underscore the importance of understanding regional imbalances in pandemic response strategies. Effectively addressing these imbalances requires accurate quantification and understanding of the regional disproportionalities of daily COVID-19 confirmed cases.

In the field of economics, various indicators have been developed to measure resource and income inequality, including one proposed by Theil incorporating information theory [12]. In reference [13], the authors demonstrate the value of using inequality indices to monitor changes in geographic inequality, and the Theil index was used to track geographic inequality over time in the COVID-19 pandemic, providing important insights to inform public health policy.

The aim of this paper is to quantify the interregional disproportionality in numbers of the confirmed cases using the Theil index, which corresponds to the Kullback-Leibler (KL) divergence in information theory [14]. It is an effective method of measuring the degree of disproportionality and objectively assessing the bias in the interregional distribution of infected individuals.

## Methods

The Theil index is commonly applied in various fields including economics, sociology, and information theory. The index quantifies the relative differences between various components of a dataset. In the context of regional analysis of the confirmed cases, the Theil index can be employed to evaluate the distribution of infected individuals across different regions. In this study, we utilize the Theil index to identify regions with disproportionate numbers of the confirmed cases relative to their population size by comparing the distribution of the confirmed cases with the overall population distribution.

The discrete form of Theil index is expressed as

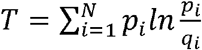

where *N* is the total number of considering regions and ln is the natural logarithm. The *p*_*i*_ which is described as the discrete probability distributions in region is the ratio of daily confirmed cases in a region and it in whole region per a day and, similarly, the *q*_*i*_ is the ratio of the population in a region and it in whole region.

The Theil index, which shares the same property as the KL divergence, is a non-symmetric metric that measures the relative entropy or difference in information represented by two distributions. It is sensitive to the interregional distribution of the confirmed cases, with its maximum value attained when the confirmed cases are concentrated in areas with the smallest population proportion. Consequently, the index tends to exhibit higher values when a small number of regions account for a large share of the confirmed cases, and conversely, lower values when the confirmed cases are more evenly distributed across regions. Notably, it remains non-negative and reaches a minimum value of 0 only when the two distributions are identical. Therefore, applying the Theil index to the timeline data of the confirmed cases, changes in the index over time can be used to quantify the degree of spread of infectious diseases and to assess whether a disproportionate concentration of infected individuals relative to the population is occurring.

## Results

We analyze the time trend of daily COVID-19 confirmed cases in the United States over 1100 days since the first reported case on January 21, 2020 [15]. Data are taken from the COVID-19 data repository at the Center for Systems Science and Engineering (CSSE) at Johns Hopkins University [16]. Population data by U.S. state were obtained from [17]. Note that population changes due to migration, births, and deaths were ignored throughout the analysis.

Before showing the results, if we use the confirmed cases as it is, the Theil index will fluctuate greatly due to the way the data is aggregated in holidays differs depending on regions, so we use the 7-day average instead of raw data.

The Fig. 1 is a two-axis graph showing the time trends of the Theil index on the left and the number of the confirmed cases on the right, on a logarithmic scale. The horizontal axis is the number of days, denoted by t in the text, elapsed since the date when the first case was reported in the U.S..

**Figure 1.**
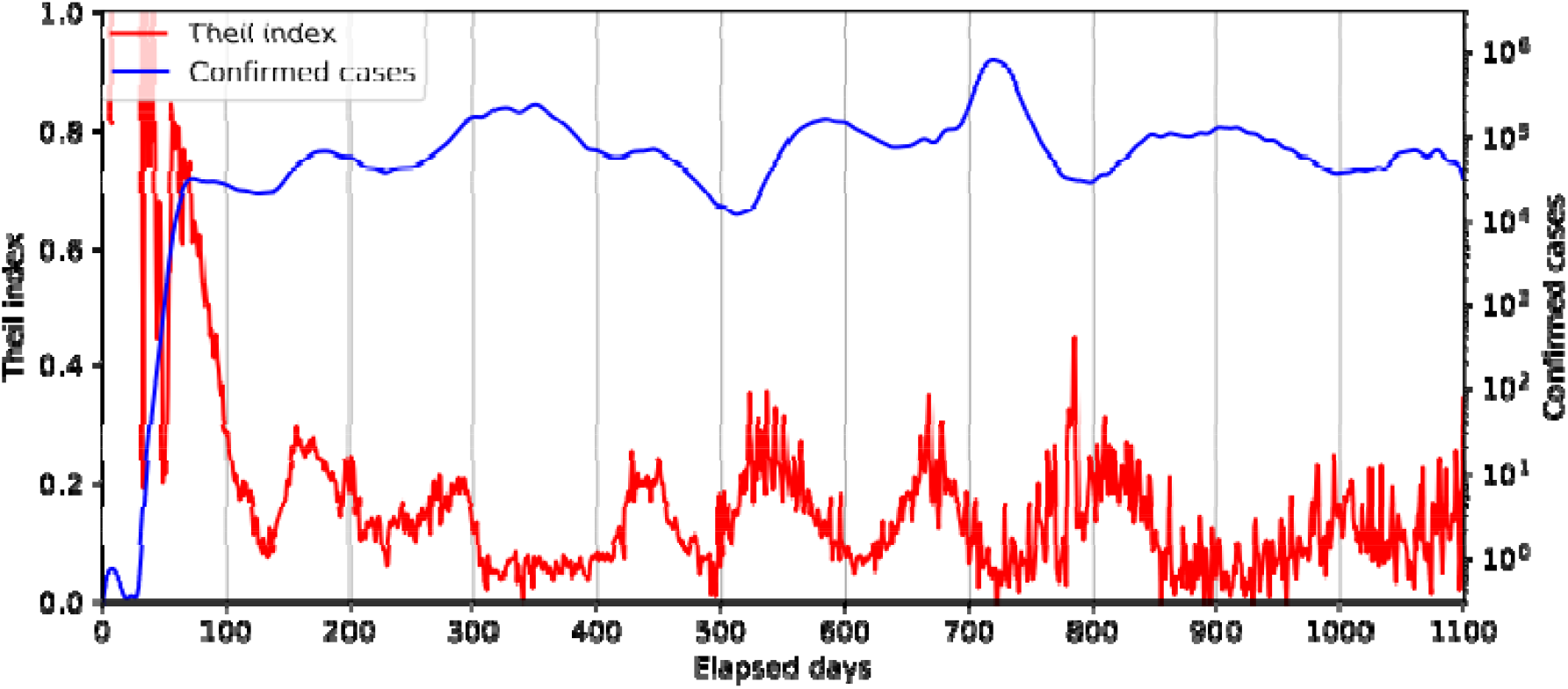
Time trends of the Theil index on the left axis and the number of 7-day averages of the confirmed cases on the right axis on a logarithmic scale are shown in the red and blue curves, respectively. The horizontal axis is the number of days elapsed since January 21, 2020.

It is important to note that increases and decreases in the Theil index simply indicate the degree of disproportionality in the confirmed cases and do not correspond to increases or decreases in the number of infected individuals. In other words, this indicator is effective when monitored in conjunction with actual trends in the number of the confirmed cases.

In Fig. 1, there are eight notable surges of the confirmed cases, occurring at around t =80(1st), 180(2nd), 350(3rd), 450(4th), 580(5th), 720(6th), 900(7th), and 1080(8th) respectively. Before the first peak, the number of the confirmed cases is quite low and the Theil index fluctuates unstably. As t increases near the 1st peak, the Theil index appears to gradually decrease, reaching a local minimum around t = 120. This implies that a fairly localized epidemic at the beginning of the COVID-19 pandemic spreads rather equally throughout the U.S.. For the other peaks in the confirmed cases, the similar phenomena can be confirmed, namely the increase of the Theil index occurs to some extent before the peak and then the Theil index decreases. This can be seen as a precursor to a surge of infected individuals as discussed in ref. [5].

It is interesting to check some examples. Practically, when the index value is high and the number of the confirmed cases is low (t = 60, 550, etc.), it indicates that the infection is occurring locally and spreading to various regions. On the other hand, when the index is low and the number of the confirmed cases is high (t = 750, etc.), it indicates that there is no clear epicenter of infection and that the number of the confirmed cases is increasing equally in different regions.

The contributions to the Theil index from each region over time are visualized by the heat-map as shown in Fig. 2 where regions with a high concentration of the confirmed cases relative to the population are colored in red, while blue regions indicate lower concentrations. There is a somewhat long interval between the deep red patches in some regions such as California, Florida and New York. In other words, the periods of intense infection represented by the deep red patches were not repeated at short intervals. This phenomenon is of great importance in infectious disease responses. Once a major epidemic in an area has subsided, the interval between subsequent outbreaks provides an opportunity to rebuild the healthcare system and implement preventive measures before the next epidemic occurs.

**Figure 2.**
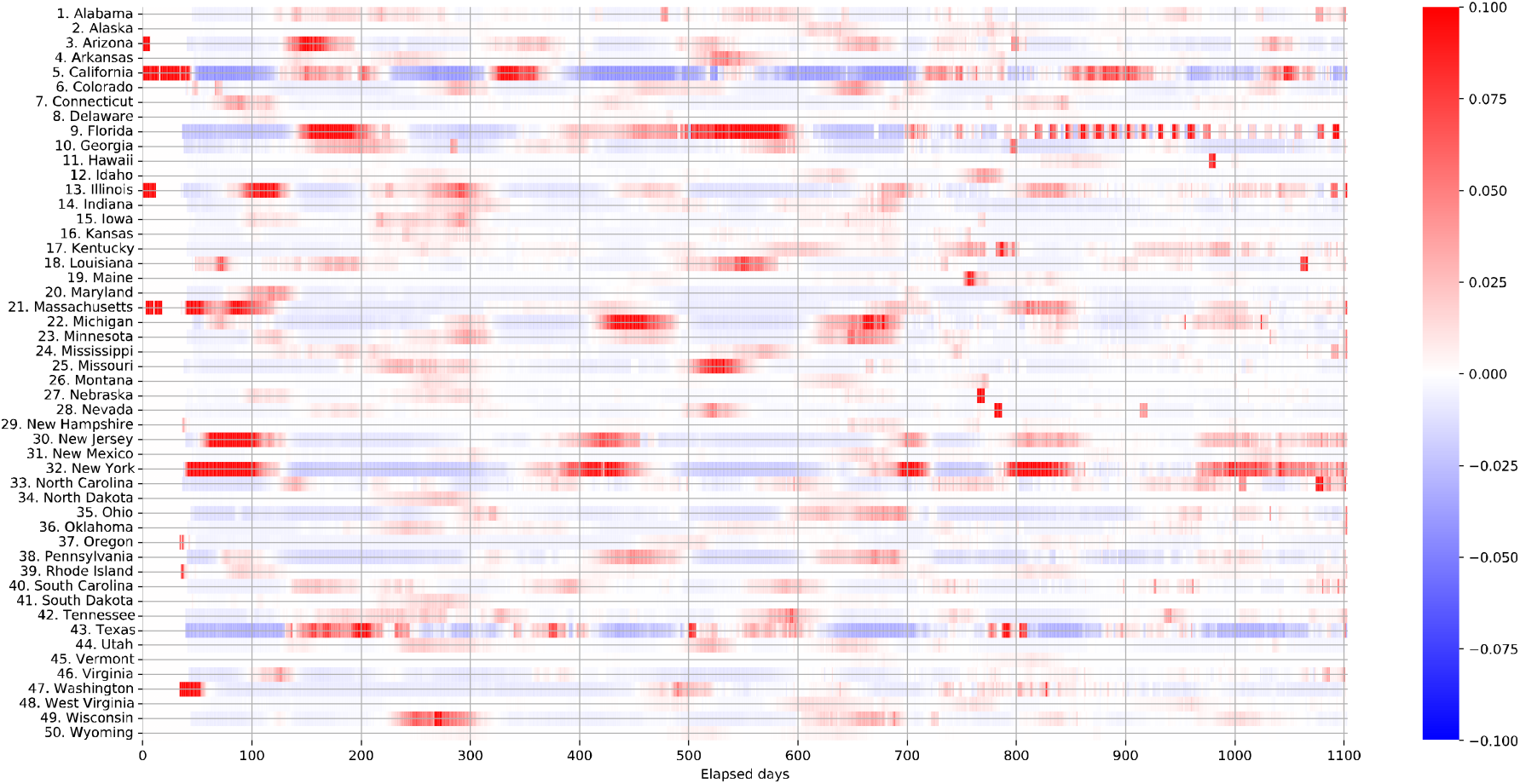
Contributions to the Theil index from each region are shown by a heat-map over the time. The positive (high concentration of prevalence) and negative (low concentration) contributions to the Theil index correspond to the deep red and blue color, respectively.

Based on the observations from Fig. 2, it is apparent that the epicenter of infectious diseases, indicated by the red patch, alternates among New York, California, and Florida. This insight is crucial for understanding the spread mechanism of future infectious diseases. Furthermore, after t = 750, both the red and blue colors fade over time, suggesting the absence of a clear epicenter and indicating a widespread outbreak of COVID-19. This suggests the ineffectiveness of countermeasures against the spread of infectious diseases under these circumstances.

## Discussion

Regional disproportionality is a critical factor influencing strategy formulation in the fight against the COVID-19 pandemic. A detailed analysis of infection patterns in different regions facilitates the development of more targeted and efficient region-specific interventions. Furthermore, if the distribution of infectious diseases is highly imbalanced, one could consider that there is an opportunity to rearrange the allocation of health care resources.

The combined monitoring of Fig. 1 and 2 allows us to show the epicenter of infectious diseases and their concentration at any time. As indicated in the results section, it was also found that once an infectious disease concentration decreases, there is some interval before the next concentration occurs. Therefore, while monitoring, it is necessary to concentrate countermeasures in areas where there is a concentration of infections and prepare for the next concentration of infections in other areas.

As it is mentioned in the previous section, enhancement of the Theil index can be seen as a precursor of a surge of the confirmed cases. In fact, just before the 1st, 4th, 6th and 7th surges, concentration of the confirmed cases is occurred in New York, and before the 2nd, 5th surges occurred in Florida.

Fig. 3(a) shows the contributions to the Theil index by region at t = 60. The horizontal axis in the figure shows the state code given in Tab. 1. There is a notable contribution to the Theil index from New York State which is prominent relative to the other regions. We also find that New Jersey and Massachusetts have a relatively large positive contribution to the Theil index due to the concentration of the number of the confirmed cases relative to the population and are better treated as the same group as New York due to their geographic proximity to each other. It can be clearly seen in Fig. 3(b) that the confirmed cases are quite localized in these regions.

**Table 1.** The list of state code used in this paper.

|  |  |  |  |  |
| --- | --- | --- | --- | --- |
| 1: Alabama | 2: Alaska | 3: Arizona | 4: Arkansas | 5: California |
| 6: Colorado | 7: Connecticut | 8: Delaware | 9: Florida | 10: Georgia |
| 11: Hawaii | 12: Idaho | 13: Illinois | 14: Indiana | 15: Iowa |
| 16: Kansas | 17: Kentucky | 18: Louisiana | 19: Maine | 20: Maryland |
| 21: Massachusetts | 22: Michigan | 23: Minnesota | 24: Mississippi | 25: Missouri |
| 26: Montana | 27: Nebraska | 28: Nevada | 29: New Hampshire | 30: New Jersey |
| 31: New Mexico | 32: New York | 33: North Carolina | 34: North Dakota | 35: Ohio |
| 36: Oklahoma | 37: Oregon | 38: Pennsylvania | 39: Rhode Island | 40: South Carolina |
| 41: South Dakota | 42: Tennessee | 43: Texas | 44: Utah | 45: Vermont |
| 46: Virginia | 47: Washington | 48: West Virginia | 49: Wisconsin | 50: Wyoming |

**Figure 3.**
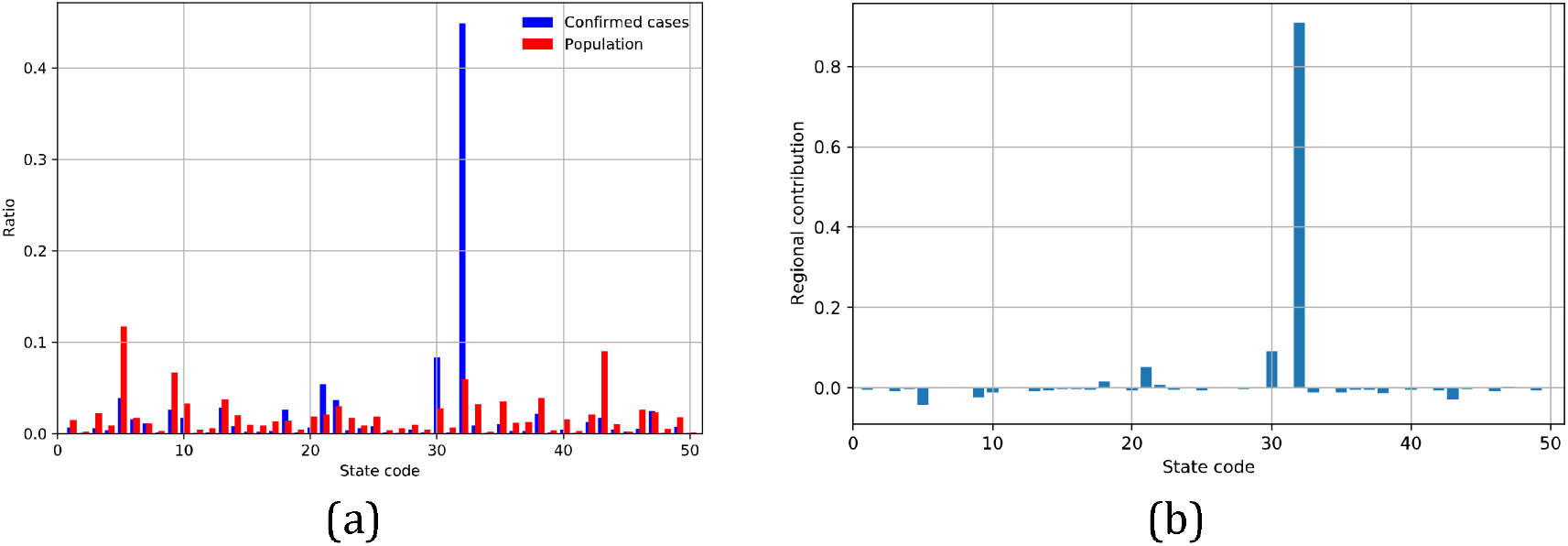
Contributions to the Theil index from each region at t = 60. The horizontal axis shows the state code given in Tab. 1. (a) Contributions to the Theil index from each region. The vertical axis shows the strength of the contribution to the Theil index. (b) Comparison of the distribution of the confirmed cases and population. The vertical axis shows the ratio of a part to the whole region for populations and for the confirmed cases.

There is a relatively large negative contribution to the Theil index from California, Florida, and Texas, which are regions of high population ratio. It is interesting to note that these are regions where there was little risk of infection at that time, but where the number of infected individuals rapidly increased after the concentration of the confirmed cases in New York took place.

The concentration of infections in New York at t = 60 as shown in Fig. 3(a) and (b) cannot be overlooked when considering infection control. The lockdown was implemented in New York city at the time when the contribution to the Theil index was concentrated in New York state. While it is impossible to assess the impact of the lockdown measures on the Theil index alone, the concentration of the confirmed cases in New York state suggests that the lockdown was implemented at the appropriate time. However, given that the contribution to the Theil index from New Jersey and Massachusetts which can be treated as the same group as New York was larger in positive values than the other regions, if lockdown measures are an appropriate response to contain COVID-19 infection, it may have been necessary to implement strong measures in these regions simultaneously to prevent the spread of COVID-19 throughout the country.

The Fig. 4 shows the contributions of the Theil index from each region at t = 550 and t = 750. At t = 550 shown in Fig. 4(a), the Theil index is a peak and trend of the confirmed cases is increasing, suggesting that the new epidemic occurred mainly in Florida and Louisiana. However, their contributions are fairly smaller than that from New York at t = 60 in Fig. 3. This indicates that the regional imbalance is much less than in the early stage of the COVID-19 pandemic. It is also interesting to look at the case at t = 750 shown in Fig. 4(b), when the confirmed cases in the US are at their maximum. In this figure, although there are several regions with large contributions to the Theil index, the epicenter of COVID-19 is no longer clear, meaning that COVID-19 is evenly distributed across the regions.

**Figure 4.**
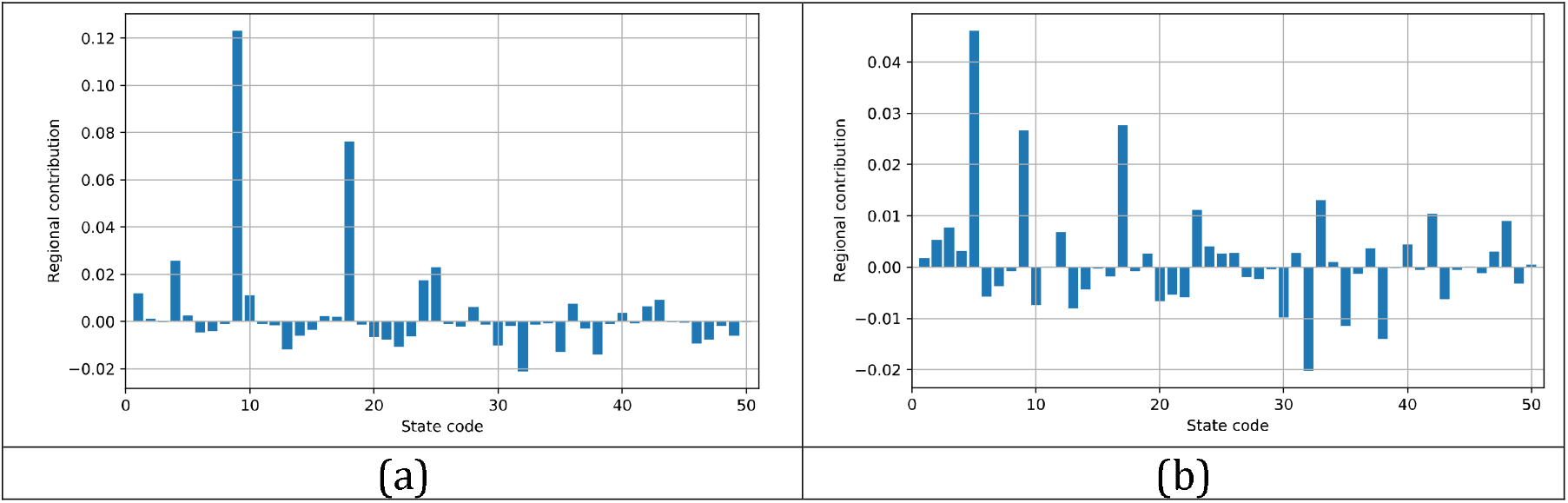
Contributions to the Theil index from each region at a specific date. The vertical axis shows the strength of contribution to the Theil index. The horizontal axis shows the state code given in Tab. 1. (a) Contributions of the Theil index at t = 550. (b) Contributions of the Theil index at t = 750.

## Conclusions

In conclusion, this study demonstrates the effectiveness of Theil index in quantifying regional disproportionalities in the confirmed cases and monitoring their evolution over time. By analyzing data of the confirmed cases in the United States, we have clarified patterns of disproportionalities in the confirmed cases, specifying epicenters and occurring localized outbreaks.

Continued monitoring and analysis of regional differences in COVID-19 transmission remains essential, especially in light of emerging variants and evolving public health responses. Our findings highlight the importance of understanding regional dynamics of infected individuals for responses of pandemic. Metrics such as Theil index provide valuable tools for policymakers and public health officials to allocate resources effectively and tailor interventions to specific regional needs.

Incorporating evidence from this study will enable policymakers to refine strategies and address the different needs of different regions, ultimately increasing the effectiveness of pandemic response efforts and mitigating the impact of future health crises.

Lastly, the decomposability of the Theil index makes it possible to quantify and compare disproportionality in groups with specific characteristics, such as age, vaccination coverage, and accessibility of health care, and identifying these disproportionalities will provide important insights for future pandemic responses.

## Data Availability

The study used ONLY openly available data. Data location is given as reference in the paper.

## Acknowledgements

We thank the members of the Division of Scientific Information and Public 184 Policy (SiPP) at the Center for Infectious Disease Education and Research (CiDER) Osaka University for useful discussions. This research was supported by The Nippon Foundation—Osaka University Project for Infectious Disease Prevention.

## Abbreviations

COVID-19: COrona Virus Infectious Disease, emerged in 2019
KL: Kullback-Leibler

